# Cachexia in Clinical Practice: Experience from an Endocrine-Led Care Model

**DOI:** 10.1101/2025.07.09.25331020

**Authors:** Anirudh Murthy, Morgan Simons, Anne Jablonski, Maurice Hurd, Alpana Shukla, Marcus D. Goncalves

## Abstract

**Background:** Cachexia is a multifactorial syndrome of involuntary weight loss, skeletal muscle wasting, and metabolic dysregulation, commonly seen in advanced cancer and other chronic diseases. Despite its prevalence and prognostic significance, effective treatment strategies remain limited, and there is no standardized model of outpatient care in the US.

**Objective:** To describe the structure, patient characteristics, and outcomes of a multidisciplinary cancer cachexia clinic embedded within an academic endocrinology practice.

**Methods:** We conducted a retrospective analysis of 103 patients referred to a single-center cachexia clinic over five years. Patients underwent comprehensive assessments including weight trajectory, nutritional status, physical performance (5x sit-to-stand test, handgrip strength), and received individualized interventions involving nutrition counseling, resistance training, and pharmacologic management.

**Results:** The median patient age was 69.7 years, with 64.1% having a cancer diagnosis (61.0% with metastases). Median monthly weight loss decreased from –0.5 kg/month in the 6 months pre-enrollment to 0.0 kg/month after 3 months post enrollment (p < 0.0001), indicating significant stabilization. The 5x sit-to-stand test improved (p = 0.022), though handgrip strength remained unchanged. Patients prescribed an exercise video program trended toward greater weight gain (β = +1.988, p = 0.079), while those prescribed protein powder tended to experience more weight loss (β = –2.102, p = 0.113), although this difference was not statistically significant.

**Conclusion:** A multimodal cachexia clinic can stabilize weight loss and improve physical function in medically complex patients. These findings support the integration of interdisciplinary approaches to cachexia management and provide a framework for evaluating future interventions in routine clinical settings.

## Introduction

Cachexia is a multifactorial syndrome characterized by involuntary weight loss, skeletal muscle wasting, and metabolic dysregulation (1). It commonly arises in the setting of chronic conditions such as heart failure, kidney disease, chronic obstructive pulmonary disease (COPD), and cancer, where the most data exists. The prevalence of cachexia in patients with advanced cancer is as high as 80% (2). Cachexia holds clinical significance and demands serious consideration beyond the changes in body weight and composition, as cachexia is associated with impairment in functional status, reduced tolerance to therapy, and increased morbidity and mortality (3–5). For example, patients with cachexia have increased complications from chemotherapy and surgery, poor therapeutic responses to immunotherapy, and worsened overall mortality, underscoring the need for focused attention on cachexia in both research and clinical practice (6,7).

Cachexia is clinically defined as unintentional body weight loss of 5% or more in 6 months or 2% or more, if the body mass index (BMI) is less than 20 kg/m^2^ (8). Unintentional weight loss can result from reductions in food intake, nutrient malabsorption, or increases in energy expenditure. Reduced food intake can be found in 40-60% of patients with advanced cancer (9). Malabsorption affects 30% to 80% of those with gastrointestinal (GI) malignancies (10), especially following radiation or surgery (11–14). Less is known about high energy expenditure. Although elevations in energy expenditure have been reported in animal models, data supporting this mechanism of cachexia in humans remains limited (15). These divergent mechanisms underscore the multifactorial nature of cachexia and the need for individualized diagnostic evaluation to inform targeted interventions.

Despite its high prevalence and important prognostic implications, the treatment of cachexia is limited by a lack of effective treatment. To date, there are no approved treatments specific to cachexia, outside of Japan, where anamorelin. a ghrelin receptor agonist, is available. In other countries, the current treatment approaches are supportive in nature and include nutritional therapy (calorie and protein goals), exercise (resistance training regimens), non-steroidal anti-inflammatory drugs (NSAIDs), steroid hormone receptor agonists (megestrol acetate and synthetic glucocorticoids), and other appetite stimulants (*e.g.*, mirtazapine, olanzapine, cannabinoids, selective serotonin reuptake inhibitors (SSRIs)) (16). While these agents can produce modest weight preservation, there is typically no improvement in functional status or survival (16). Overall, there remains no consensus approach to care for patients with cachexia in day-to-day clinical practice.

Several institutions have developed dedicated cachexia clinical care programs to address this unmet need. These programs integrate pharmacologic symptom management, nutritional counseling, and physical rehabilitation, and have demonstrated improvements in symptom burden, appetite, emotional well-being, and quality of life despite limited changes in weight or lean mass (17–21). Importantly, outcomes from these clinics highlight the feasibility and value of coordinated, individualized care in stabilizing functional decline and supporting patients and caregivers in the face of progressive disease (16,21,22). However, real-world implementation remains variable, and few data are available from U.S.-based endocrinology-led models of care or settings outside palliative care and oncology. Here, we describe the structure, patient population, and outcomes of a multidisciplinary cancer cachexia clinic integrated into an academic endocrinology division, offering insight into how cachexia care can be delivered in diverse practice environments.

## Methods

Data were collected through a manual review of the electronic medical record (EMR) for a 5-year period between July 1, 2018, and July 31, 2023, focused on patients referred to Endocrinology for care of unintentional weight loss, cachexia, protein-calorie malnutrition, poor appetite, anorexia, sarcopenia, or muscle weakness. The study was reviewed by the Weill Cornell Medicine IRB and determined to qualify for exemption per the Code of Federal Regulations on the Protection of Human Subjects (45 CFR 46), under category 4(ii).

### Clinic design and approach

All clinical care was provided in the Endocrinology outpatient practice at Weill Cornell Medicine, located in New York City, NY, USA. Patients were evaluated and treated by two providers, an American Board of Internal Medicine (ABIM)-certified endocrinologist (MDG) and a nurse practitioner (AFJ) with expertise in endocrinology, obesity medicine, and reproductive health. Both were licensed to practice in New York State. Local providers were encouraged to refer patients for evaluation of unintentional weight loss if they experienced a weight loss of more than 5% over 6 months due to any suspected cause. At the time of presentation, a detailed body weight history was obtained, including self-reported recall of young adult weight, peak body weight and age, and weight values from 12, 6, and 3 months ago. All available body weight measurements were extracted from the EMR and discussed with the patient to identify discrepancies and errors. A GI review of systems was performed with specific inquiries into the presence or absence of anorexia, odynophagia, dysphagia, dysguesia, early satiety, abdominal pain, nausea, vomiting, diarrhea, constipation, and steatorrhea. The patient was asked about the number and size of meals, including a description of the types and amount of food consumed at each meal.

As part of the clinical evaluation, all patients underwent a focused physical examination to assess skeletal muscle mass, strength, and the size of subcutaneous adipose tissue reserves. Visual inspection and palpation were used to evaluate muscle wasting, as well as fat loss. Muscle strength was graded on the standard 5-point Medical Research Council (MRC) scale in the deltoids, biceps, triceps, hip flexors, knee flexors, and knee extensors. Handgrip strength was measured using a Jamar dynamometer, and averaged over three trials with the dominant hand. When feasible, lower extremity performance was assessed using either the five-times sit-to-stand (5xSST) or the 30-second sit-to-stand (30sSST) test, selected based on safety and functional capacity (23,24).

EMR-based laboratory values were reviewed to rule out reversible causes of weight loss, including hypercalcemia, hyperglycemia, hyponatremia, uremia, hypogonadism, adrenal insufficiency, and hyperthyroidism. Abnormal results were further evaluated and treated as per standard clinical practice. Laboratory data were also evaluated for evidence of malnutrition, including iron, folate, and vitamin B12 deficiency using complete blood counts and red cell indices, as well as vitamin D deficiency. When recent laboratory values were unavailable in the EMR, the tests were requisitioned. Inflammation was assessed using C-reactive protein (CRP) and white blood cell differential, including neutrophil and lymphocyte counts.

Nutritional counseling was provided to ensure adequate caloric and protein intake, with targets of >30 kcal/kg/day and >1.0 g/kg/day of protein, adjusted based on individual weight, comorbidities, and intake history. A sample 3-day meal plan was provided to each patient, designed to meet these goals using familiar and culturally appropriate foods. Dietary recommendations emphasized energy-dense, protein-rich meals and snacks. When food intake alone was insufficient to meet protein targets, supplementation with a whey protein isolate (20-30 g) was recommended with or without creatine monohydrate (5 g per day). A daily multivitamin was recommended to help address potential micronutrient deficiencies and support overall nutritional adequacy. For patients who continued to lose weight after two weeks on the initial diet plan, caloric intake was increased by 300 kcal per day, with adjustments made incrementally until weight stabilization or gain was achieved.

After one month of treatment, patients who were unable to comply with the dietary plan were offered pharmacologic appetite stimulation. Low-dose mirtazapine was preferred when sleep disturbance was present, olanzapine when nausea was present, and a SSRI in cases of concurrent depressed mood. Steroid hormone receptor agonists (megestrol acetate and glucocorticoids) were avoided, given the high rate of deleterious adverse events, particularly on the endocrine system (16). In some patients, anabolic-androgenic steroids (testosterone, oxandrolone) were used.

Patients were encouraged to begin a home-based resistance training program three days per week, consisting of a brief warm-up followed by lunges, wall push-ups, squats, overhead presses, bicep curls, and bent-over rows. A set of links were provided to online videos demonstrating each movement. Patients were instructed to perform these exercises using available household items such as hand weights, resistance bands, or water bottles to provide resistance, and to complete 3-4 sets of 10 repetitions each.

### Statistical Analysis

We summarized baseline demographics, symptom characteristics, and results using descriptive statistics. Because the data were not normally distributed, we used medians and interquartile ranges to describe the continuous data, as well as frequencies for categorical data.

As noted previously, cachexia is a progressive condition for which there is no curative treatment. To that end, we determined that the best method to investigate the effectiveness of our intervention was to compare the rates of weight change before and after intervention, as even a decrease in rate of weight loss would represent a meaningful improvement in some patients. The rate of weight change was calculated by dividing the difference in weight by the time interval between visits, multiplied by a hundred. Wilcoxon signed-rank test was used to compare the rates of weight change between the three time intervals (i.e. 12 months before intervention to 6 months before intervention (interval I), 6 months before intervention to initiation of intervention (interval II), and initiation of intervention to 3 months after intervention (interval III)). Bonferroni correction was applied to compute adjusted p-values. For functional parameters, Wilcoxon signed-rank test was used to determine the significance of the change between baseline and three months after intervention. A regression analysis was performed to assess the relationship between the change in weight during interval III and the specific interventions. Spearman’s rank correlation coefficient was used to evaluate the correlation between the rate of weight change in interval III and various biomarkers at baseline. All analysis and graphing were performed using R version 4.3.3 and Microsoft Excel.

## Results

### Patient demographics and characteristics

A total of 103 patients were treated. Most identified as white (65%) and male (53.4%). The median age was 69.7 years (IQR: 56.2 - 75.7 years), and 53.9% had a normal body mass index (BMI) (18.5 - 24.9 kg/m^2^) at the time of initial visit. Most patients had diagnosed cancer (64.1%). Of the patients with cancer, the most common types were lung (17.1%), pancreatic (12.9%), esophageal (11.4%), head and neck (5.7%), or prostate (5.7%). Most patients with cancer had diagnosed metastases (61.0%). Comorbidities included diabetes (24%) and thyroid disease (30%) (Table 1).

**Table 1.** Patient Demographics.

| Parameters | N | % |
| --- | --- | --- |
| **Race** |  |  |
| White | 67 | 65.0 |
| African American | 10 | 9.7 |
| Asian | 3 | 2.9 |
| Other | 12 | 11.7 |
| Declined | 11 | 10.7 |
| **Sex** |  |  |
| Male | 55 | 53.4 |
| Female | 48 | 46.6 |
| **BMI (kg/m<sup>2</sup>)** |  |  |
| Underweight [$<18.5$] | 25 | 24.5 |
| Normal Weight [$18.5$-$24.9$] | 55 | 53.9 |
| Overweight [$25$-$29.9$] | 19 | 18.6 |
| Obese [$>30$] | 3 | 2.9 |
| **Cancer** |  |  |
| Yes | 66 | 64.1 |
| No | 37 | 35.9 |
| **Metastasis** |  |  |
| Yes | 36 | 61.0 |
| No | 23 | 39.0 |
| **Cancer type** |  |  |
| Lung | 12 | 17.1 |
| Pancreatic | 9 | 12.9 |
| Esophageal | 8 | 11.4 |
| Head and Neck | 4 | 5.7 |
| Prostate | 4 | 5.7 |
| Lymphoma | 3 | 4.3 |
| Leukemia | 3 | 4.3 |
| Colorectal | 3 | 4.3 |
| Upper GI | 3 | 4.3 |
| Neuroendocrine | 3 | 4.3 |
| Breast | 3 | 4.3 |
| Urothelial | 2 | 2.9 |
| Glioblastoma | 2 | 2.9 |
| Thyroid | 2 | 2.9 |
| Renal | 2 | 2.9 |
| Other | 7 | 10.0 |
| **Diabetes** |  |  |
| Yes | 24 | 23.3 |
| No | 79 | 76.7 |
| **Thyroid Disease** |  |  |
| Yes | 30 | 29.1 |
| No | 73 | 70.9 |

Symptoms of reduced food intake were common and frequently accompanied by objective evidence of impaired muscle function. Poor appetite and/or reduced food intake were reported in 44 (42.7%) patients. Of the 36 participants assessed for baseline hand grip strength, 17 (47%) demonstrated low grip strength (25). Among the 29 participants with recorded 5x sit-to-stand (SST) times, 23 (79%) performed below standard thresholds (26–28).

To better understand systemic factors that may contribute to these symptoms and the broader cachexia phenotype, we assessed baseline hormonal, inflammatory, and nutritional biomarkers in the entire cohort. The median baseline testosterone level among males was 457 ng/dL [IQR: 267.5–564 ng/dL], with 21% meeting criteria for hypogonadism (testosterone <264 ng/dL) (29). The median baseline TSH was 2.0 mIU/L [IQR: 1.3–3.0 mIU/L], and 4% of participants were found to have hyperthyroidism (TSH <0.4 mIU/L) (30). The median hs-CRP concentration was 5.2 mg/L [IQR: 1.3–14.4 mg/L], with 57% demonstrating elevated levels (>3 mg/L) (31,32). The median IL-6 level was 4.5 pg/mL [IQR: 2-5 pg/mL], and 8% of individuals had elevated levels of IL-6 (> 7pg/mL) (33,34). The median neutrophil-lymphocyte ratio (NLR) was 3.28 [IQR: 2.1– 5.4], and 43% had an elevated NLR (>3.83) (35–37). Median albumin was 3.8 g/dL [IQR: 3.4–4.2 g/dL], with hypoalbuminemia (<3.5 g/dL) observed in 26% of participants (38).

Based on the clinical evaluation, providers identified contributing etiologies of weight loss and assigned specific ICD diagnoses accordingly, which guided individualized intervention strategies (Table 2). The most frequently documented ICD diagnoses included abnormal weight loss (R63.4, 67.0%), cachexia (R64.0, 45.6%), and anorexia (R63.0, 34.0%). The most common interventions included recommendations to increase protein and calories (76.7%), use of protein powder with or without creatine monohydrate (66.0%), and structured exercise videos (60.2%). A minority of patients received appetite stimulation therapy (29.1%), including olanzapine or mirtazapine, anabolic androgenic steroids (18.4%) like testosterone or oxandrolone, and/or pancreatic enzyme replacement (12.6%) for malabsorption.

**Table 2.** Diagnoses and Interventions.

|  | N | % |
| --- | --- | --- |
| **Diagnosis (ICD code)** |  |  |
| Abnormal weight loss (R63.4) | 69 | 67.0 |
| Cachexia (R64.0) | 47 | 45.6 |
| Anorexia (R63.0) | 35 | 34.0 |
| Sarcopenia (M62.84) | 27 | 26.2 |
| Moderate protein-calorie malnutrition (E44.0) | 20 | 19.4 |
| Unspecified protein-calorie malnutrition (E46.0) | 12 | 11.7 |
| Feeding difficulties (R63.3) | 2 | 1.9 |
| **Intervention (Yes)** |  |  |
| Protein/Calorie increase | 79 | 76.7 |
| Protein Powder (w/ and w/o creatine) | 68 | 66.0 |
| Exercise Videos | 62 | 60.2 |
| Appetite Stimulant | 30 | 29.1 |
| Anabolic Steroids/Hormone Therapy | 19 | 18.4 |
| Pancreatic Enzyme Replacement | 13 | 12.6 |

Fifty-two (50%) patients did not return for a second visit, while 18 (17%), 13 (13%), and 10 (19%) returned for 1, 2, or more follow-up appointments.

### Weight trajectory over time

Median patient weight was 62.9 kg [IQR: 52.8 - 76.0 kg] twelve months prior to clinical entry, 65.0 kg [IQR: 53.25 - 79.45 kg] six months before entering the clinic, and 60.3 kg [IQR: 49.7 - 74.25 kg] at the initial visit (baseline). Three months after baseline, the median weight remained relatively unchanged (59.2 kg [IQR: 48.5 - 71.9 kg]) (Figure 1a).

**Figure 1:**
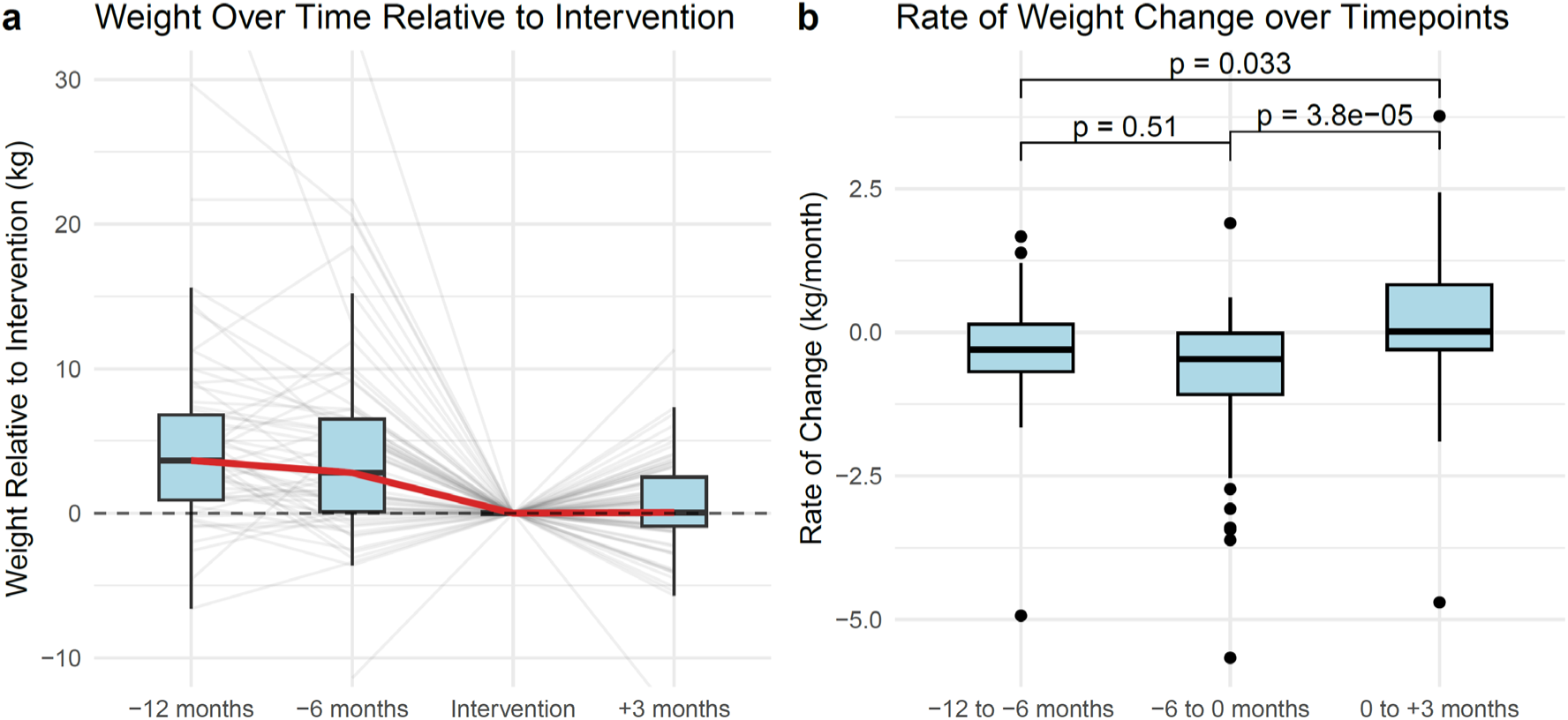
Weight Change Over Time. (a) *Weight changes over time relative to the intervention.* Box plots depict weight (kg) relative to the time of intervention at four time points: 12 months prior, 6 months prior, the intervention time point (reference), and 3 months post-intervention (N=103). Each thin gray line represents an individual participant’s trajectory. The thick red line shows the group median trend over time. (b) *Rate of weight change over consecutive time intervals.* Box plots show the distribution of individual rates of weight change (kg per month) across three intervals: −12 to −6 months, −6 to 0 months (intervention), and 0 to +3 months post-intervention (N=47). Pairwise comparisons between intervals were assessed using Wilcoxon signed-rank test, with p-values indicated above brackets.

Over interval I, patients lost a median of 0.3 kg/month [IQR: -0.7 - 0.1 kg/month]; over interval II, patients lost a median of 0.5 kg/month [IQR: -1.1 - 0.0 kg/month], and over interval III, patients’ median weight was unchanged (0.0 kg/month [IQR: -0.3 - 0.8 kg/month]) (Figure 1b). Wilcoxon signed-rank analysis of the monthly rate of weight change between the intervals showed a statistically significant difference between interval II and interval III (p < 0.0001); likewise, there was a modest difference between interval I and interval III (p = 0.033). As expected, there was no significant difference between interval I and interval II (Table 3).

**Table 3.** Pairwise Comparison of Rate of Weight Change.

| Comparison | N (Pairs) | Raw p-value | Adjusted p-value* |
| --- | --- | --- | --- |
| -12 to -6 months vs. -6 months to 0 months | 47 | 0.171 | 0.513 |
| -6 to 0 months vs. 0 to 3 months | 47 | 1.25e-5 | 0.000038 |
| -12 to -6 months vs. 0 to 3 months | 47 | 0.011 | 0.033 |

\* Bonferroni correction for multiple comparisons

### Effect of intervention on weight change

A regression analysis was performed to assess the relationship between the change in weight during interval III and the specific interventions. Participation in exercise videos was associated with the greatest weight gain (β = +1.988), with a trend towards significance (p = 0.079). On the other hand, use of protein powder (with or without creatine) was associated with the most weight loss (β = -2.1020), although this was not statistically significant (p = 0.113) (Table 4).

**Table 4.** Multiple Linear Regression Results: Effects of Intervention on Weight Change.

| Variable | Coefficient (B) | Standard Error | t-value | p-value |
| --- | --- | --- | --- | --- |
| **Intercept** | -0.325 | 0.886 | -0.367 | 0.715 |
| Exercise Videos | +1.988 | 1.114 | 1.784 | 0.079<sup>†</sup> |
| Protein Powder (w/ or w/o Creatine) | -2.120 | 1.323 | -1.603 | 0.113 |
| Protein/Calorie Increase | +1.060 | 1.275 | 0.831 | 0.409 |
| Anabolic Steroids/Hormone Therapy | -0.123 | 1.018 | -0.121 | 0.904 |
| Appetite Stimulant | +0.593 | 0.877 | 0.676 | 0.501 |
| Creon | -0.012 | 1.195 | -0.010 | 0.992 |

<sup>†</sup> $p < 0.1$

### Association of initial biomarkers to weight change in interval III

There was a weak linear relationship between the percentage of weight change over interval III and initial hs-CRP level (R^2^=0.17, *P=0.051*); however, no association between weight change and serum IL-6 (R^2^=0.023, *P=0.48*) (Supplementary Figure 1). No associations were found between weight change and other hematologic or endocrine parameters (Supplementary Figures 2 and 3).

### Effect of intervention on functional status

There was a significant improvement in the 5xSST performance (p = 0.042) during interval III (Figure 2a) from 15.7 seconds [IQR: 11.5 - 19.7 sec] at baseline to 11.1 [IQR: 10.2 - 18.7 sec] after 3 months. There was no difference (30 kg [IQR: 18 – 38 kg] to 27 kg [IQR: 20.5 - 33.5 kg]) in strength as assessed by hand grip strength during interval III (p = 0.714) (Figure 2b).

**Figure 2:**
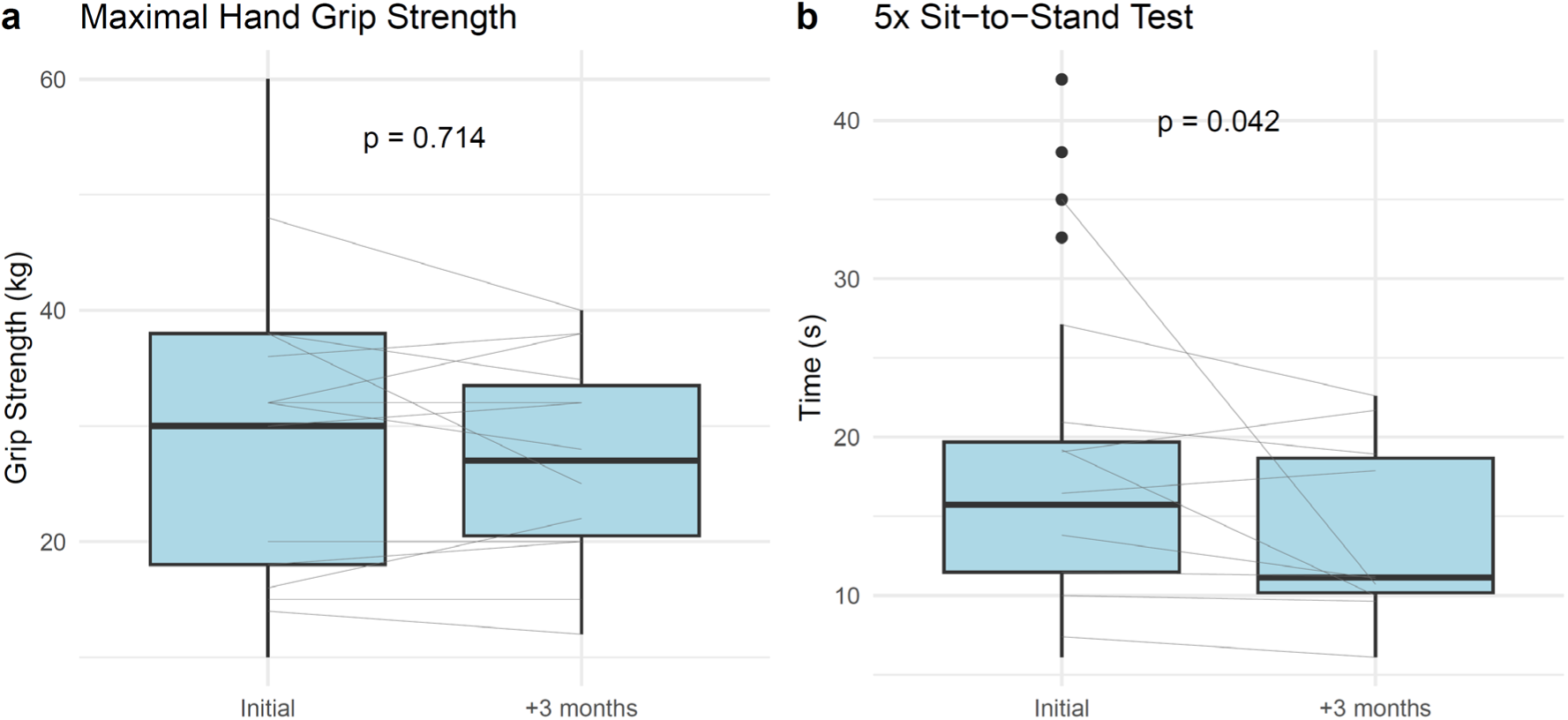
Change in Performance and Strength Over Time. (a) *Performance on the 5x sit-to-stand test before and after 3 months.* Box plots display the distribution of times (seconds) required to complete the 5x sit-to-stand test at the initial assessment and at 3 months (N=36). Thin gray lines connect paired measurements for individual participants. The p-value reflects the statistical comparison of test performance between time points, showing a trend towards significance. Lower times indicate improved functional performance. (b) *Hand grip strength at baseline and after 3 months.* Box plots illustrate maximal hand grip strength (kg) measured at the initial assessment and at 3-month follow-up (N=29). Each thin gray line represents the paired measurements for an individual participant. The p-value denotes the result of the Wilcoxon signed-rank testing between time points, showing no significance.

## Discussion

Here we report data from a single-center, endocrinology-led cancer cachexia clinic embedded within an academic outpatient practice. In a disease marked by progressive decline and limited treatment options, our findings offer evidence that structured, multimodal care can stabilize body weight and improve physical performance over a short time frame.

In line with prior studies (17–21), we observed that the median weight of patients was stabilized following enrollment in the clinic. While weight gain was not universal, there was statistically significant deceleration in weight loss trajectory, from a median of -0.5 kg/month prior to enrollment to 0.0 kg/month after three months. Notably, these outcomes were achieved in a medically complex cohort with heterogeneous causes of weight loss, including a substantial proportion of older adults with metastatic cancer. These data help to define the current standard of care and establish a baseline against which novel agents can be compared. For instance, ponsegromab, a novel monoclonal antibody against GDF-15, was recently shown to increase weight by 0.7 kg/month over 3 months in phase II trials with associated improvements in appetite, symptoms, and functional status in patients with cancer cachexia (39). Based on our data, this change is clinically meaningful and a significant advance over the current standard of care.

Previous work has shown that hand grip strength and sit-to-stand times are prognostic indicators in patients with cachexia (40,41). We found that the stabilization of body weight in our population was accompanied by a significant improvement in physical performance (SST times), but not strength (hand grip). These results may suggest that patients may have experienced improvements in mobility or neuromuscular coordination, rather than gains in pure muscle strength, which may require longer-term or higher-intensity interventions. Alternatively, the handgrip test may not be sensitive enough to capture changes in this time frame, as others have mentioned (42). Nevertheless, maintenance of hand grip strength, as opposed to loss, may in and of itself represent a favorable outcome in a condition where no standard of care exists.

Among the specific interventions, the use of a home exercise prescription demonstrated the strongest association with weight gain (β = +1.988, *p* = 0.079), though this did not reach statistical significance. The evidence of exercise, either alone or in combination with other modalities of treatment, remains mixed. Exercise therapy shows mechanistic evidence for mitigating cancer cachexia by preserving muscle mass, reducing inflammation, and improving metabolic function (43,44). However, current human clinical evidence shows no consistent improvements in lean mass, strength, or quality of life (45).

In contrast, patients receiving protein powder supplementation with or without creatine experienced greater weight loss (β = -2.102) in multivariate testing. Increased protein intake, above 1.0-2.0 g/kg/day, is recommended to help mitigate muscle depletion, with some evidence suggesting that specific amino acids, such as leucine, may enhance muscle protein synthesis (46). However, in clinical trials, there is no robust evidence that protein supplementation improves outcomes (16). A plausible explanation may be the increased satiety of protein powder compared to carbohydrates and fats. This effect is partially mediated by modulation of appetite-regulating hormones such as ghrelin, cholecystokinin, glucagon-like peptide-1, and peptide YY (47). The impact of protein supplementation on subsequent energy intake and long-term appetite regulation requires further study in larger sample sizes.

This study has important limitations. First, the absence of a control group precludes causal inference. Patients represented a heterogeneous population with several underlying etiologies for weight loss, diverse tumor types, and treatment regimens. Importantly, we did not assess their disease burden or the management of their underlying condition, which may influence weight dynamics. Referral patterns may have introduced selection bias. For example, the patients sent to our clinic may have had higher baseline engagement with care or access to better support services. Roughly half of the patients only had one clinic visit, limiting longitudinal assessment. Additionally, body composition was not measured, so it remains unclear whether stabilized weight reflected preservation of lean mass, fat mass, or fluid shifts.

Despite these limitations, this study presents a real-world model for outpatient cachexia care that is both feasible and effective in altering clinical trajectories. By integrating detailed nutritional counseling, strength-based physical therapy, and targeted pharmacologic support into a single visit, we achieved clinically meaningful stabilization in a population with otherwise poor expected outcomes. As emerging therapeutics for cachexia continue to be tested, data such as ours establish a critical foundation for interpreting their clinical relevance in everyday practice. In a field that has long-lacked effective options, even modest gains in function or stabilization of decline may represent significant progress.

## Data Availability

All data produced in the present study are available upon reasonable request to the authors

## Acknowledgments

None

---

**Supplementary Figure 1:**
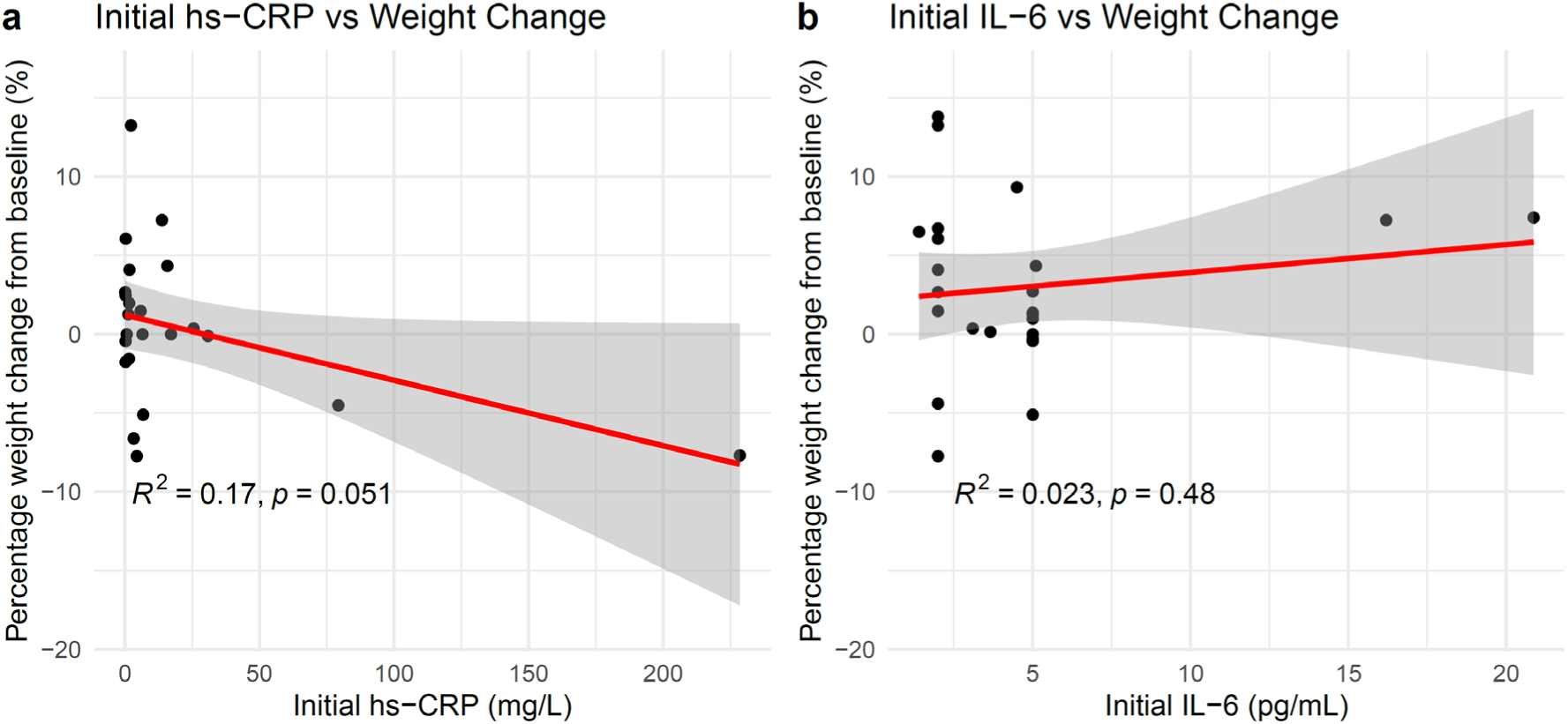
Associations between baseline inflammatory biomarkers and percentage weight change. Scatter plots show the relationships between (a) baseline hs-CRP (N=28) and (b) baseline IL-6 levels (N=25) with percentage weight change from baseline over the study period. Each point represents an individual participant. Red lines indicate linear regression fits, with shaded areas depicting 95% confidence intervals. Baseline hs-CRP showed a non-significant trend toward greater weight loss (R² = 0.17, *p* = 0.051), while no significant association was observed between baseline IL-6 and weight change (R² = 0.023, *p* = 0.48) using Spearman’s rank correlation coefficient.

**Supplementary Figure 2:**
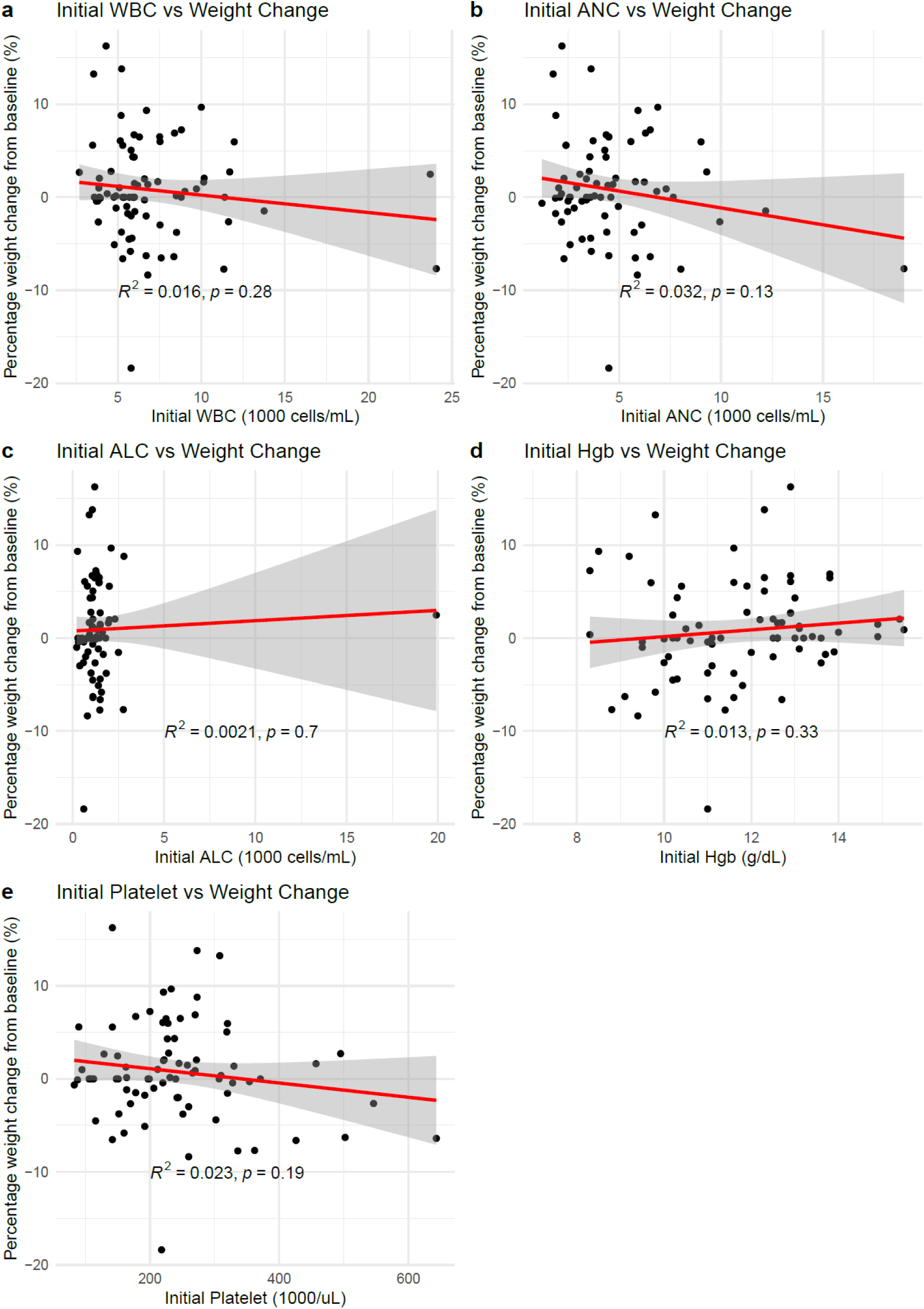
Associations between baseline hematologic parameters and percentage weight change. Scatter plots illustrate the relationships between baseline laboratory values and subsequent percentage change in body weight over the study period: (a) white blood cell count (WBC) (N=95), (b) absolute neutrophil count (ANC) (N=84), (c) absolute lymphocyte count (ALC) (N=83), (d) hemoglobin (Hgb) (N=95), and (e) platelet count (N=95). Each point represents an individual participant. Red lines indicate linear regression fits, and shaded regions show 95% confidence intervals. No statistically significant associations were observed between any hematologic parameter and weight change using Spearman’s rank correlation coefficient. R² and *p*-values for each regression are displayed within each panel.

**Supplementary Figure 3:**
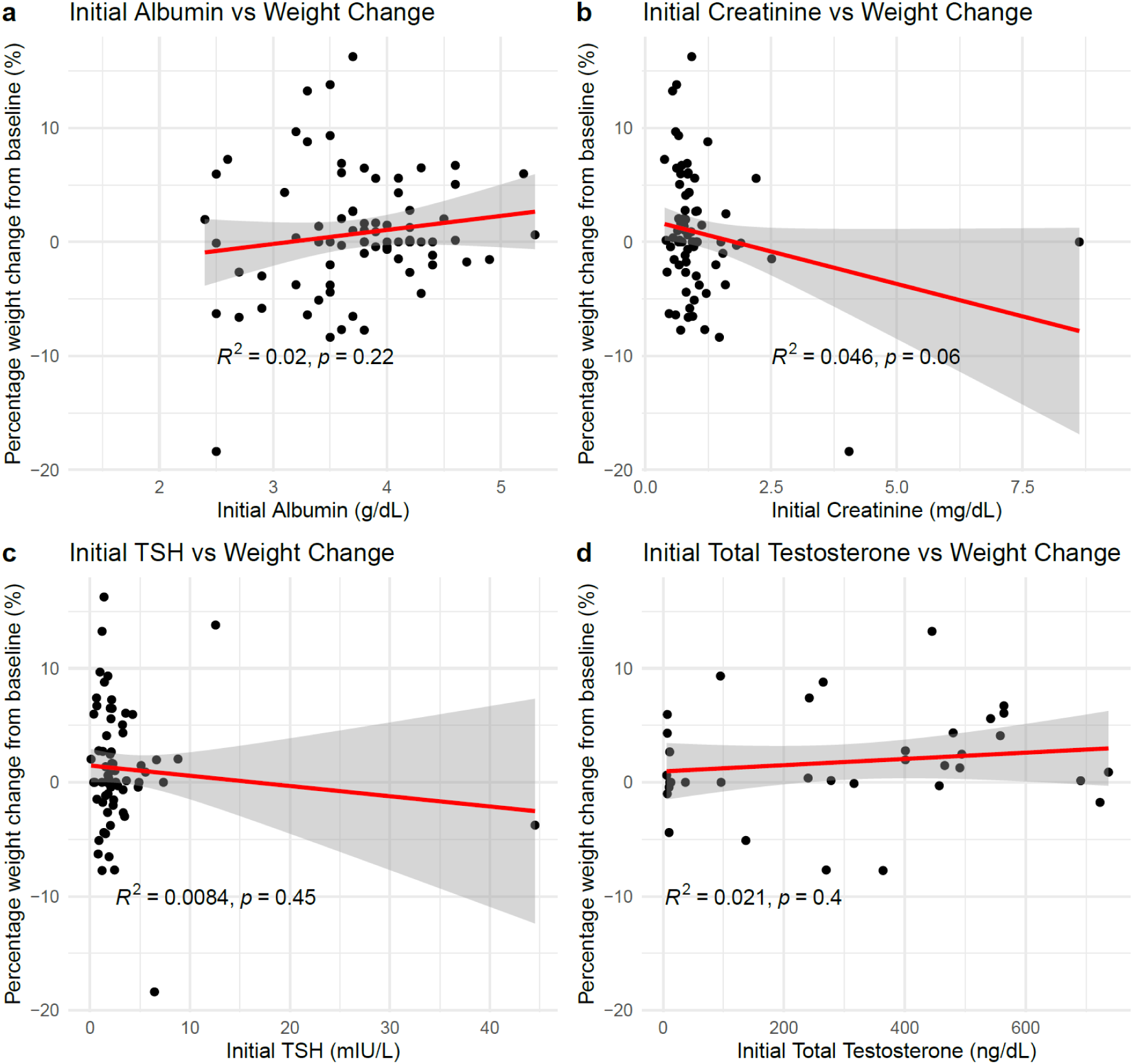
Associations between baseline metabolic and hormonal markers and percentage weight change. Scatter plots depict the relationships between initial laboratory values and percentage change in body weight over the study period: (a) serum albumin (N=96), (b) serum creatinine (98), (c) thyroid-stimulating hormone (TSH) (N=82), and (d) total testosterone (N=41). Each point represents an individual participant. Red lines indicate linear regression fits, and shaded areas show 95% confidence intervals. No statistically significant associations were observed between these biomarkers and weight change using Spearman’s rank correlation coefficient. R² and *p*-values for each regression are reported within each panel.

